# Pattern of Severity of Road Traffic Injuries Among Pedestrians in Low- and Middle-Income Countries: A Systematic Review

**DOI:** 10.1101/2021.02.13.21251689

**Authors:** Neeraj Sharma, Mohan Bairwa, S. D. Gupta, D. K. Mangal

## Abstract

**Background:** Low-and middle-income countries (LMICs) contribute about 93 per cent of road traffic injuries (RTIs) and deaths worldwide with a significant proportion of pedestrians (22 per cent). Various scales are used to assess the pattern of injury severity, which are useful in predicting the outcomes of RTIs. We conducted this systematic review to determine the pattern of RTI severity among pedestrians in LMICs.

**Methods:** We searched the electronic databases PubMed, CINHAL, CENTRAL, Web of Science, Scopus, EMBASE, ProQuest and SciELO, and examined the references of the selected studies. Original research articles published on the RTI severity among pedestrians in LMICs during 1997-2016 were eligible for this review. Quality of publications was assessed using an adapted Newcastle-Ottawa Scale of observational studies. Findings of this study were presented as a meta-summary.

**Results:** Five articles from 3 LMICs were eligible for the systematic review. Abbreviated Injury Score, Glasgow Coma Scale and Maxillofacial Injury Severity Score were used to assess the injury severity in the selected studies. In a multicentric study from China (2013), 21, 38 and 19 per cent pedestrians with head injuries had AIS scores 1-2, 3-4 and 5-6, respectively. In another study from China (2010), the proportion of AIS score 1-2 and AIS score 3 and above (serious to un-survivable) injuries occurred due to crash with sedan cars were 65 and 35 per cent, respectively. Such injuries due to minivan crashes were 49.5 per cent and 50.5 per cent, respectively. Two studies Ikeja, Nigeria (2014) and Elazig, Turkey (2009) presented, 24.5 and 32.5 per cent injured had a severe head injury (GCS < 8), respectively. In another study from Ibadan, Nigeria (2014), the severe maxillofacial injuries were seen in the victims of car/minibus pedestrian crashes 46 per cent, and 17 per cent had a fatal outcome.

**Conclusion:** A varied percent of pedestrians (24.5 to 57 percent) had road traffic injuries of serious to fatal nature, depending on type of collision and injury severity scale. This study pressed the need to conduct studies with a robust methodology on the pattern of RTI severity among pedestrians to guide the programme managers, researchers and policymakers in LMICs to formulate the policies and programmes to save the pedestrian lives.

**African relevance:**

- Prior RTI research reveals that pedestrians and cyclists were at the highest risk of fatality of in Sub-Saharan Africa, whereas motorcyclists had significantly higher fatality rates in Asian countries such as Malaysia and Thailand (1–3).
- Fifty-seven type of injury severity scoring systems have been developed to assess the injury severity for triage and timely decision making for patient treatment need, outcome prediction, quality of trauma care, and epidemiological research and evaluation (4,5).
- We found two studies from sub-Saharan Africa in this review which showed that severe pedestrian injuries ranged from 24.5 to 46 per cent of total pedestrian RTIs.
- Despite the findings of review affected by limited and variegated sample, it could be useful to guide for future research.

## BACKGROUND

Road traffic injury (RTI) is a neglected and challenging global health issue especially among low- and middle-income countries (LMICs). Globally, RTIs resulted in 1.35 million deaths in 2018 and LMICs contributed about 93 per cent of total RTI deaths, although, having only 60 per cent of the total registered vehicles (6). RTI is a leading cause of disease burden in one of the most productive age group of 15-29 years (6,7). In LMICs, the estimated cost of RTIs peaked up to 3 per cent of total GDP (1), and RTI has emerged in an epidemic proportion (8).

About half of RTI deaths are among the vulnerable road users, namely, pedestrians, cyclists, and motorcyclists; predominantly due to pedestrian-vehicle collisions (1). In 2015, pedestrian road injuries contributed to about 24.5 million DALYs globally among all age groups (7). Pedestrians contributed to a large share (39 per cent) among RTI deaths in 2017 (9). In pedestrian-vehicle collisions, the traumatic brain injuries were the commonest cause of death (2). Pedestrians and cyclists were at the highest risk of death in Sub-Saharan Africa, whereas motorcyclists had significantly higher fatality rates in Asian countries such as Malaysia and Thailand (2). Industrialization, economic growth, and subsequently, increased numbers and speed of vehicles may have an impact on the pattern of the RTI severity among pedestrians (3,10).

Fifty-seven type of injury severity scoring systems have been developed and used for triage and timely decision making for patient treatment need, outcome prediction, quality of trauma care and epidemiological research and evaluation (4,5). Some commonly used injury severity scoring systems are (a) Anatomical scoring systems [Abbreviated injury scale (AIS), Injury severity score (ISS), International Classification of Diseases 9 based injury severity score (ICISS), New injury severity score (NISS), Anatomical Profile, Anatomical Index]; (b) Physiological scoring systems [Glasgow coma scale (GCS), Trauma score, Revised trauma score, Prognostic Index, Acute Trauma Index, Triage Index]; and (c) Combined scoring systems [A severity characteristics of trauma (ASCOT), Trauma and injury severity score (TRISS), Harborview Assessment of Risk of Mortality] (11,12).

These injury severity scores had been used for decades to estimate the injury severity and for the prediction of outcomes among victims of RTIs in LMICs (4). Evidence on the pattern of injury severity among pedestrians will be useful for better prediction and subsequently planning of trauma care and improving the quality of care. Hence, this review was conducted to synthesize the evidence on the pattern of RTI severity among pedestrians in LMICs.

## METHODS

### Protocol and Registration

This systematic review is reported in accordance with the Meta-analysis of Observational Studies in Epidemiology (MOOSE) (Additional File – 1) checklist and retrospectively registered after preliminary searches completed in the International Prospective Register of Systematic Reviews (PROSPERO - CRD42017073853). Available from: http://www.crd.york.ac.uk/PROSPERO/display_record.php?ID=CRD42017073853

### Information Sources

We searched PubMed, Web of Science, Scopus, EMBASE, ProQuest, CINHAL, CENTRAL, and SciELO databases for relevant studies. Initial 20 pages of Google Scholar were also reviewed to supplement the search results. We contacted the authors for further information on injury severity scale and pedestrian injuries data which were not available in the selected articles, however, no author has responded to the queries sent through emails.

### Search strategy

We listed indexing terms and other keywords to describe concept clusters. The searches were performed on the titles and abstracts. The initial search covered the keywords ‘Road Traffic Accident/Injury/Crash’ AND ‘Low and Middle-Income Countries’. Then, the search was restricted for the duration from 1 January 1997 to 31 December 2016. The final search was conducted on 1^st^ September 2017. This search strategy was modified according to the need of search engines (Additional file – 2).

We used the World Bank list of low and middle-income countries to identify and shortlist the publications of LMICs (13) (Additional file – 3). Rayyan software® was used to remove duplication among search records based on titles and abstracts (14). Reference lists of selected articles were examined for additional data sources; however, it did not provide new article.

### Inclusion and exclusion criteria

#### Study design

All original studies were included providing data on the pattern of injury severity.

#### Study population and outcome

The population of interest was pedestrians from all age groups those who were affected by road traffic injuries. The outcome of interest was the pattern of the severity of road traffic injuries.

#### Report characteristics

Eligible studies were limited to peer-reviewed articles in all languages from LMICs only. Editorials, letter to the editor, errata, book chapter, commentaries and study protocols were excluded.

## Data management and extraction

We imported all identified references to Rayyan software^®^ and combined them into a single library to remove the duplicate records. Then first author (NS) screened all the articles by titles and abstracts for inclusion or exclusion of the articles. Second author-verified all the search results and screened the studies. A PRISMA flow diagram of the article’s selection procedure was prepared (Figure 1).

**Figure 1:**
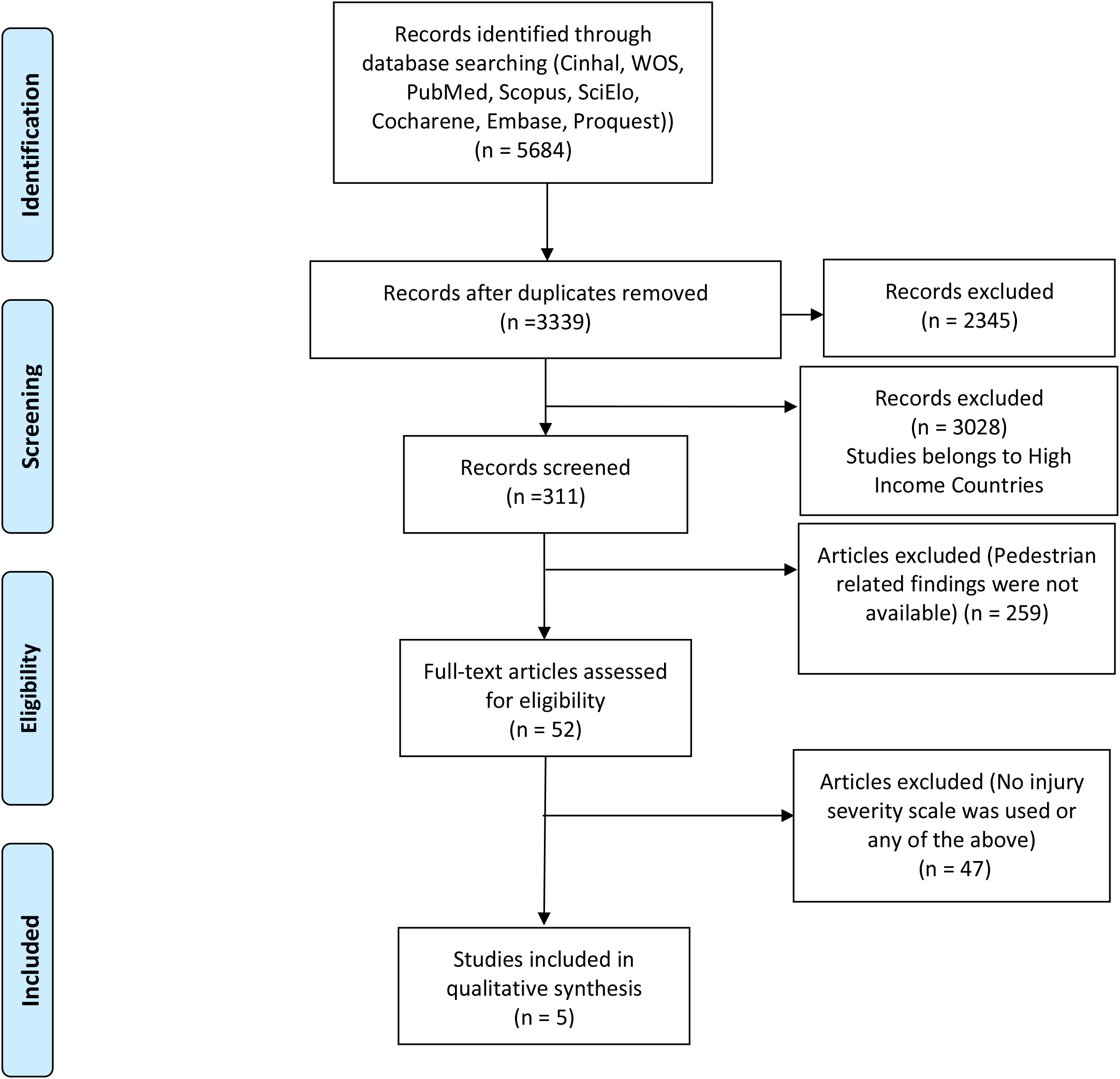
PRISMA Flow Diagram *From:* Moher D, Liberati A, Tetzlaff J, Altman DG, The PRISMA Group (2009). *P*referred *R*eporting *I*tems for *S*ystematic Reviews and *M*eta-*A*nalyses: The PRISMA Statement. PLoS Med 6(7): e1000097. doi:10.1371/journal. pmed1000097 **For more information, visit www.prisma-statement.org**.

In next step, the first authors (NS) performed the full-text review of all shortlisted studies and extracted the relevant data from all finally included studies (Additional file – 4). This was cross-checked by the second reviewer (MB) to assess and ensure the correctness of data extraction.

Any inconsistencies between the reviewers were sorted out by consensus, or arbitration by the review team (DKM and SDG). The data extraction from the eligible studies included the name of the author(s), year of publication, study location, study setting, study design, sampling method, severity scale, participants characteristics, sample size, and key findings.

### Quality assessment

The quality of studies was assessed using an adapted version of the Newcastle-Ottawa Scale (Additional file – 5). In this scale, 4 criteria are related to selection (maximum 5 points), 1 criteria for comparability (maximum 2 points), and 2 criteria related to outcome (maximum 3 points). All seven criteria of the scale in studies with primary data (cross-sectional studies) and six criteria in studies with secondary data (retrospective studies) were identified. A maximum of 10 scores (ranged 1 to 10) were assigned to the studies conducted on the primary data. While a maximum 9 scores (ranged 1 to 9) were assigned studies with secondary data analysis. Non-response criteria were not assessed in the studies based on secondary data.

### Data synthesis

We were unable to conduct a meta-analysis due to high methodological variability, hence, meta-summary (15) was used to aggregate the findings from all the studies.

## RESULTS

Five studies met the eligibility criteria in this systematic review. A summary of the characteristics of the selected articles is shown in ***Additional file - 6***. All included studies were cross-sectional. Three studies were based on primary data (16–18) and two were based on secondary data sources (19,20). Of the total, 2 studies were carried out in Nigeria (16,17), 2 in China (18,20) and 1 in Turkey (19). Three studies included pedestrian from all the age groups (16,19,20) and two studies had specific age groups – 15 years and below (17) and 16 years and above (18). Three studies were population-based (17,18,20) and two were hospital-based (16,19). These studies used following injury severity scales: **AIS** (18,20), **GCS** (17,19) and **MFISS** (16).

### AIS

Studies by Nie Jin et al (2010) and Zhao et al (2013) used AIS to assess the severity of injury in population-based settings in China. This scale was designed to classify the RTIs affected persons. Although, it did not intend to provide an outcome prediction of RTIs. The latest version of AIS (AIS-90) has 1312 codes of both blunt and penetrating injury types. It divides the body into six regions (head/neck, face, chest, abdominal/pelvic contents, extremities/pelvic girdle, external) and scores the severity of injuries for each region. The scores are classified into six categories based on the severity level: AIS 1-Minor, AIS 2-Moderate, AIS 3-Serious, AIS 4-Severe, AIS 5-Critical, AIS 6-Unsurvivable (11,21,22).

### GCS

Solagberu et al (2014) in Nigeria and Tokdemir et al (2009) in Turkey used GCS to assess the severity of injury during the hospitalization (17,19). It is useful in predicting the outcomes after head injury. The GCS scores are based on the motor, verbal, and eye-opening response; each domain is scored independently within 1-5 points depends upon the victim’s response. Then, all three section’s individual scores were summed up to estimate the GCS score of the injured (23,24). The GCS scores of 3–8, 9–12 and 13–15 denote severe, moderate, and mild head injury, respectively.

### MFISS

Aladelusi et al (2014) used MFISS to assess the severity among pedestrian RTIs involving maxillofacial injuries of all age groups in a hospital setting in Nigeria (16). The MFISS method includes the 3 highest AIS scores of the maxillofacial region, then it is combined with the Injury Severity Scores (ISS) for 3 maxillofacial functional parameters, namely, malocclusion, limited mouth opening, and facial deformity (25). The ISS is a sum of the squares of the highest AIS score in each of the three most severely injured body regions (26).

### Pattern of severity based on Injury Severity Scales

Included studies were of poor quality. The studies did not provide sufficient information on the representativeness of the sample, adequacy of sample size and non-response. In addition, the study participants varied by different characteristics, hence, we were unable to pool the available data (Additional file – 7).

Nie Jin et al (2010) analyzed the data of the pedestrians injured due to crashes with sedan and minivans (20). The study found:

- The proportion of minor to moderate (AIS score 1 and 2) and serious to un-survivable (AIS score 3 and above) injuries due to sedan cars were 65 and 35 per cent, respectively.
- Minor to moderate and serious to un-survivable injuries due to minivans crashes were 49.5 per cent and 50.5 per cent, respectively.

Zhao et al (2013) analyzed data on all type of car crashes (18) and found:

- Pedestrians had 21, 38 and 19 per cent head injuries with AIS 1-2, 3-4 and 5-6 scores, respectively. 42.5 per cent of pedestrians RTIs were fatal.

Assessment of severity based on AIS revealed that 35 to 57 per cent of pedestrian road traffic injuries involving head were serious in nature. It may have been varied due to different age groups, type of vehicular occupants, and body parts involved in the accidents.

Solagberu et al (2014) analyzed the data of 108 child-pedestrians aged ≤ 15 years with head injuries and found: About 37 per cent had a mild head injury (GCS >13); 30.5 per cent had a moderate head injury (GCS 9-12), while 32.5 per cent had a severe head injury (GCS < 8) (17).

Tokdemir et al (2009) analyzed data on head injuries among all age groups and found: About 60 per cent had a mild injury (GCS ≥ 13); 15.8 per cent had a moderate injury (GCS 9-12), while 24.5 per cent had a severe injury (GCS of ≤ 8) (27).

However, this category belonged to head injuries only, 24.5 to 32.5 per cent affected by severe injuries based on GCS. In children, higher proportion of head injuries were severe in nature.

Aladelusi et al (2014) analyzed the data on maxillo-facial injuries among pedestrians and found (16) that the median MFISS score of victims of car/minibus was 9 and motorcycle collision was 4. The most severe maxillofacial injury was seen in the victim of car/minibus pedestrian crashes 46 per cent, and 17 per cent had a fatal outcome.

Use of different scales may also affected the range of severity of injuries in the study participants, in addition to the factors described. Still, we could say that about one forth to more than half of injuries were severe/ serious in nature.

## DISCUSSION

This systematic review assessed the pattern of the severity of pedestrian RTIs in LMICs, which is a specific area of pedestrian RTI research. We found 5 studies from 3 LMICs in the last two decades. These studies used only three injury severity assessment scales (AIS, GCS, and MFISS). Serious/ severe injuries variably ranged from 24.5 per cent (head injuries of all type) to 57 per cent (car crashes involving head injuries) of the total injuries in the included studies. Fatal injuries ranged from 17 per cent (maxillofacial injuries in crash with minibus/ cars) to 42.5 per cent (head injuries in car crashes). We did not find any common pattern of the severity of injuries among affected pedestrians across studies due to the heterogeneous nature of evidence available. The reasons of heterogeneity were participant characteristics such as age groups, type of injuries, different study settings, classification used to access the injury severity, and varying target population which restricted us to combine the study findings.

Pedestrian injuries belong mainly to vulnerable sections of society including children, elderly, poor, laborers’ etc. (28,29), which further increases their vulnerability due to poor health, subsequently, disability and deaths. The injury severity scales significantly contributed to saving the lives of injury victims (11,30). An abundance of literature is available on the use of injury severity assessment scales among pedestrian RTIs in high-income countries (31–34). Their use could help to save the loss of pedestrian lives due to RTIs and improve pedestrian RTI management in LMICs. However, the applicability of these evidence from high-income countries may need tailoring in LMICs because of differences in culture and education, developmental stage, inadequate infrastructure, and poor implementation of road safety laws. This possible gap could be filled in by well-designed research for utilization of injury severity scores to improve treatment outcomes of pedestrian RTIs. Although, healthcare providers in LMICs also face several constraints in the implementation of the injury severity scores, in terms of inadequate infrastructure, manpower, poor training, and capacity building (11). This further incapacitates them to deliver better quality trauma care to vulnerable road users such as pedestrians. This is high time for researchers and policymakers in LMICs to increase the research focus on the use of injury severity assessment scales among pedestrian RTIs.

Although, available studies suffered from poor quality and the reasons for poor quality including but not limited to inadequate sample size, lack of details on non-response and insufficient statistical analysis. This indicates the need to increase awareness, generate more evidence on the utility of injury severity scales for prediction of the outcome and prognosis of the injuries, and develop consensus for the use of injury severity scales specifically targeted to pedestrian RTIs among program managers, healthcare providers and researchers. It may be helpful to provide a sound body of evidence for use of the scales among pedestrian RTIs to improve trauma care. It will enable healthcare providers to predict outcomes more judiciously and prioritize the care of injured persons, and program managers and policy planners to escalate and develop preventive measures for pedestrian road traffic injuries.

The major limitation of this systematic review was the heterogeneity among the included studies, for example, data sources (primary and secondary data), study settings (hospital-based, registry-based, police record based, and population-based), outcomes measured, heterogenous participant profiles, and most importantly different types of the injury severity scales. Although, this was the first attempt in form of a systematic review to explore the pattern of the injury severity pattern of RTI affected pedestrians among LMICs, which calls for more research and evidence to reduce the disease burden of RTI among pedestrians in LMICs.

## CONCLUSION

A variable segment of pedestrian RTIs (from 1/4^th^ to more than ½) were of severe/ serious nature. Results of the studies were variable; hence, we were unable to reach at a conclusion about the pattern of the severity of injuries. Despite the pedestrians being the vulnerable road users, there was a lack of depth on the assessment of the severity of road traffic injuries among pedestrians. More research is required to assess and launch new initiatives on use of injury severity scores for identifying the severity pattern and predicting the outcomes of care. This will help strengthen and formulate the policies and programmes to address this neglected area of the RTI research in LMICs.

## Data Availability

All data generated or analyzed during this study are included in this published article [and its supplementary information files].

## LIST OF ABBREVIATIONS

AIS: Abbreviated Injury Scale
ASCOT: A Severity Characteristics of Trauma
CINHAL: Cumulative Index to Nursing and Allied Health Literature
DALYs: Disability Adjusted Life Years
EMBASE: Excerpta Medica dataBASE
GCS: Glasgow Coma Scale
GDP: Gross Domestic Products
ICISS: International Classification of Diseases 9 based injury severity score
ISS: Injury Severity Score
LMICs: Low- and Middle-Income Countries
MFISS: Maxillo-Facial Injury Severity Score
MOOSE: Meta-analysis Of Observational Studies in Epidemiology
NISS: New Injury Severity Score
PRISMA: Preferred Reporting Items for Systematic Reviews and Meta-Analyses
PROSPERO: Prospective Register of Systematic Reviews
RTI: Road Traffic Injury
SciELO: Scientific Electronic Library Online
TRISS: Trauma and Injury Severity Score

## DECLARATIONS

### Ethics approval and consent to participate

This systematic review does not require ethical clearance as it used secondary data which is already in the public domain. In addition, this study has not used any unit-level data.

### Consent for publication

Not applicable

### Author’s Contribution

NS and MB contributed to the design, analysis, interpretation, and drafting of the manuscript. DKM and SDG contributed to the interpretation and drafting of the manuscript. All authors have reviewed the manuscript critically for important intellectual content and have given final approval of the version to be published.

## Acknowledgements

This paper was a part of MPH-Capstone (JHSPH, USA and IIHMR University, Jaipur) of the first author (NS).

## Legends

Additional file – 1: MOOSE Checklist

Additional file – 2: Search Strategy

Additional file – 3: World Bank list of low and middle-income countries

Additional file – 4: List of articles excluded after full-text review

Additional file – 5: Adapted Newcastle-Ottawa Scale adapted for cross-sectional studies

Additional file – 6: Summary of studies reported severity pattern of road traffic injuries among pedestrians in LMICs

Additional file – 7: Quality Appraisal of shortlisted studies with Newcastle-Ottawa Scale

## REFERENCES

1. World Health Organization. Global status report on road safety [Internet]. Injury prevention. 2015. Available from: http://www.who.int/violence_injury_prevention/road_safety_status/2015/en/

2. Nnadi MON, Bankole OB, Fente BG. Etiology and treatment outcome of pedestrians with traumatic brain injuries from road traffic crashes. Int J Med Res Heal Sci [Internet]. 2015;4(4):861–4. Available from: https://www.ijmrhs.com/medical-research/etiology-and-treatment-outcome-of-pedestrians-with-traumatic-brain-injuries-from-road-traffic-crashes.pdf

3. UNIDO. Industrialization in Africa and Least Developed countries [Internet]. 2016. Available from: https://www.unido.org/sites/default/files/2016-09/G20_new_UNIDO_report_industrialization_in_Africa_and_LDCs_0.pdf

4. Mehmood A, Hung YW, He H, Ali S, Bachani AM. Performance of injury severity measures in trauma research: A literature review and validation analysis of studies from low-income and middle-income countries. BMJ Open. 2019;9(1):1–11.

5. Palmer C. Major trauma and the injury severity score--where should we set the bar? Annu Proc Assoc Adv Automot Med [Internet]. 2007;51:13–29. Available from: http://www.ncbi.nlm.nih.gov/pubmed/18184482%5Cnhttp://www.pubmedcentral.nih.gov/articlerender.fcgi?artid=PMC3217501http://www.ncbi.nlm.nih.gov/pubmed/18184482%5Cnhttp://www.pubmedcentral.nih.gov/articlerender.fcgi?artid=PMC3217501

6. WHO. Global Status Report on Road [Internet]. 2018. Available from: https://www.who.int/violence_injury_prevention/road_safety_status/2018/English-Summary-GSRRS2018.pdf

7. Kassebaum NJ, Arora M, Barber RM, Bhutta ZA, Brown J, Carter A, et al. Global, regional, and national disability-adjusted life-years (DALYs) for 315 diseases and injuries and healthy life expectancy (HALE), 1990-2015: a systematic analysis for the Global Burden of Disease Study 2015. Lancet [Internet]. 2016 Oct 8;388(10053):1603–58. Available from: https://search.proquest.com/docview/1828270592?accountid=136944

8. Mandal BK, Yadav, Nath B. Pattern and distribution of pedestrian injuries in fatal road traffic accidental cases in Dharan, Nepal. J Nat Sci Biol Med [Internet]. 2014;5(2):320–4. Available from: https://www.ncbi.nlm.nih.gov/pmc/articles/PMC4121907/?report=reader#!po=4.16667

9. Ritchie H, Roser M. Causes of Death [Internet]. Our World in Data. 2019 [cited 2019 Sep 19]. Available from: https://ourworldindata.org/causes-of-death

10. Watanabe R, Katsuhara T, Miyazaki H, Yasuki T. Research of the relationship of pedestrian injury to collision speed, car-type, impact location and pedestrian sizes using human FE model (THUMS Version 4). Stapp Car Crash J [Internet]. 2012;56:269–321. Available from: http://www.embase.com/search/results?subaction=viewrecord&from=export&id=L370163854

11. Kim YJ. Injury severity scoring systems: a review of application to practice. Nurs Crit Care [Internet]. 2012;17(3):138–50. Available from: http://doi.wiley.com/10.1111/j.1478-5153.2012.00498.x

12. Chawda MN, Hildebrand F, Pape HC, Giannoudis P V. Predicting outcome after multiple trauma: which scoring system? Injury [Internet]. 2004 Apr [cited 2017 Oct 1];35(4):347– 58. Available from: http://linkinghub.elsevier.com/retrieve/pii/S0020138303001402

13. World Bank Country and Lending Groups – World Bank Data Help Desk [Internet]. [cited 2017 Sep 27]. Available from: https://datahelpdesk.worldbank.org/knowledgebase/articles/906519-world-bank-country-and-lending-groups

14. Ouzzani M, Hammady H, Fedorowicz Z, Elmagarmid A. Rayyan - a web and mobile app for systematic reviews. Syst Rev [Internet]. 2016;5(1):210. Available from: http://dx.doi.org/10.1186/s13643-016-0384-4

15. Sandelowski M, Barroso J, Voils CI. Using Qualitative Metasummary to Synthesize Qualitative and Quantitative Descriptive Findings. Res Nurs Heal [Internet]. 2007;30(1):99–111. Available from: https://www.ncbi.nlm.nih.gov/pmc/articles/PMC2329806/pdf/nihms45489.pdf

16. Aladelusi TO, Akinmoladun I V, Olusanya OO, Akadiri OA, Fasola AO. Evaluation of pedestrian road traffic maxillofacial injuries in a Nigerian tertiary hospital. Afr J Med Med Sci [Internet]. 2014;43(4):353–9. Available from: https://www.scopus.com/inward/record.uri?eid=2-s2.0-84941138399&partnerID=40&md5=e40e9dfac948217328b86d713bdc3542

17. Solagberu BA, Osuoji RI, Ibrahim NA, Oludara MA, Balogun RA, Ajani AO, et al. Child pedestrian injury and fatality in a developing country. Pediatr Surg Int [Internet]. 2014 Jun;30(6):625–32. Available from: https://link.springer.com/article/10.1007%2Fs00383-014-3516-8

18. Zhao H, Yang G, Zhu F, Jin X, Begeman P, Yin Z, et al. An investigation on the head injuries of adult pedestrians by passenger cars in China. Traffic Inj Prev [Internet]. 2013 Nov;14(7):712–7. Available from: http://search.ebscohost.com/login.aspx?direct=true&db=rzh&AN=107909774&site=ehost-live&scope=site

19. Tokdemir M, Kafadar H, Turkoglu A, Deveci SE, Colak C, M. T, et al. Comparison of the severity of traumatic brain injuries in pedestrians and occupants of motor vehicles admitted to firat health center: A five-year series in an Eastern Turkish city. Med Sci Monit [Internet]. 2009;15(1):PI1–4. Available from: https://www.medscimonit.com/download/index/idArt/869506

20. Nie J, Yang J, Li F. A study on pedestrian injuries based on minivan and sedan real-world accidents. In: International Conference on Optoelectronics and Image Processing [Internet]. Newyork: IEEE; 2010. Available from: https://ieeexplore.ieee.org/document/5663269

21. Barnes J, Hassan A, Cuerden R, Cookson R, Banbury J. Comparison of injury severity between AIS 2005 and AIS 1990 in a large injury database. Ann Adv Automot Med [Internet]. [cited 2017 Oct 1];53:5–7. Available from: https://dspace.lboro.ac.uk/2134/12111

22. Cambridge Orthopedics. Abbreviated injury scale [Internet]. [cited 2017 Oct 1]. Available from: http://www.cambridgeorthopaedics.com/easytrauma/classification/trauma scores/abbreviated_injury_scale.htm

23. Teasdale G, Jennett B. Assessment of coma and impaired consciousness. Lancet [Internet]. 1974 Jul [cited 2017 Oct 1];304(7872):81–4. Available from: http://linkinghub.elsevier.com/retrieve/pii/S0140673674916390

24. Teasdale G, Jennett B. Assessment and prognosis of coma after head injury. Acta Neurochir (Wien) [Internet]. 1976 Mar [cited 2017 Oct 1];34(1–4):45–55. Available from: http://link.springer.com/10.1007/BF01405862

25. Zhang J, Zhang Y, El-Maaytah M, Ma L, Liu L, Zhou LD. Maxillofacial Injury Severity Score: Proposal of a new scoring system. Int J Oral Maxillofac Surg [Internet]. 2006;35(2):109–14. Available from: https://www.ijoms.com/article/S0901-5027(05)00242-0/fulltext

26. World Health Organization. Global strategy on human resources for health: Workforce 2030. Who [Internet]. 2016;64. Available from: http://apps.who.int/iris/bitstream/10665/250368/1/9789241511131-eng.pdf?ua=1%5Cnhttp://www.who.int/hrh/resources/pub_globstrathrh-2030/en/http://apps.who.int/iris/bitstream/10665/250368/1/9789241511131-eng.pdf?ua=1%5Cnhttp://www.who.int/hrh/resources/pub_globstrathrh-2030/en/

27. Tokdemir M, Kafadar H, Turkoglu A, Deveci SE, Colak C. Comparison of the severity of traumatic brain injuries in pedestrians and occupants of motor vehicles admitted to firat health center: A five-year series in an Eastern Turkish city. Med Sci Monit [Internet]. 2009 Jan;15(1):PI1–4. Available from: https://www.medscimonit.com/download/index/idArt/869506

28. Fokam PG, Njock R. International Journal of Surgery Open Injury patterns in road traf fi c victims comparing road user categories□: Analysis of 811 consecutive cases in the emergency department of a level I institution in a. Int J Surg Open [Internet]. 2018;10:30–6. Available from: https://reader.elsevier.com/reader/sd/pii/S2405857217300748?token=76C48A39DE116216DC98E76EEA81EA66161A5F7492E383CAE2E2200F9220D832D7CBF66A89D694DE447BB439535A41A3

29. Lee JS, Kim YH, Yun JS, Jung SE, Chae J, Chung M. Characteristics of patients injured in road traffic accidents according to the new injury severity score. Ann Rehabil Med [Internet]. 2016;40(2):288–93. Available from: https://www.e-arm.org/journal/view.php?doi=10.5535/arm.2016.40.2.288

30. Yang J, Yao J, Otte D. Correlation of different impact conditions to the injury severity of pedestrians in real world accidents. 19th Int Tech Conf Enhanc Saf Veh [Internet]. 2005;1–8 Paper No. 05-0352. Available from: https://pdfs.semanticscholar.org/e6ba/1daa20064457ad3ddc746b9a37a482983123.pdf

31. Charters KE, Gabbe BJ, Mitra B. Population incidence of pedestrian traffic injury in high-income countries: A systematic review. Injury [Internet]. 2017;48(7):1331–8. Available from: http://dx.doi.org/10.1016/j.injury.2017.05.021

32. Elurua N, Bhata CR, Hensher DA. A mixed generalized ordered response model for examining pedestrian and bicyclist injury severity level in traffic crashes. Accid Anal Prev [Internet]. 2008;40(3):1033–54. Available from: https://doi.org/10.1016/j.aap.2007.11.010

33. Kim J-K, Ulfarsson GF, Shankar VN, Mannering FL. A note on modeling pedestrian-injury severity in motor-vehicle crashes with the mixed logit model. Accid Anal Prev [Internet]. 2010 Nov [cited 2018 Jul 16];42(6):1751–8. Available from: http://linkinghub.elsevier.com/retrieve/pii/S0001457510001326

34. Lee C, Abdel-Aty M. Comprehensive analysis of vehicle–pedestrian crashes at intersections in Florida. Accid Anal Prev [Internet]. 2005 Jul [cited 2018 Jul 16];37(4):775–86. Available from: http://linkinghub.elsevier.com/retrieve/pii/S0001457505000564

