## Supplemental Data 1 for "Pattern of Severity of Road Traffic Injuries Among Pedestrians in Low- and Middle-Income Countries: A Systematic Review"

**MOOSE Guidelines for Meta-Analyses and Systematic Reviews of Observational Studies***

| | Topic | Page numbe |
|---|---|---|
| *Title* | Identify the study as a meta-analysis (or systematic review) | 1 |
| *Abstract* | Use the journal's structured format | 1 |
| *Introduction* | **Present:** | 2 |
| | The clinical problem | 3 |
| | The hypothesis | NA |
| | A statement of objectives that includes the study population, the condition of interest, the exposure or intervention, and the outcome(s) considered | 3 |
| *Sources* | **Describe:** | |
| | Qualifications of searchers (eg, librarians and investigators) | 3-4 |
| | Search strategy, including time period included in the synthesis and keywords | 4 |
| | Effort to include all available studies, including contact with authors | 4 |
| | Databases and registries searched | 3 |
| | Search software used, name and version, including special features used (e.g. explosion) | 4 |
| | Use of hand searching (e.g, reference lists of obtained articles) | 4 |
| | List of citations located and those excluded, including justification | 5 |
| | Method of addressing articles published in languages other than English | NA |
| | Method of handling abstracts and unpublished studies | NA |
| | Description of any contact with authors | 5 |
| *Study Selection* | **Describe** | |
| | Types of study designs considered | 4 |
| | Relevance or appropriateness of studies gathered for assessing the hypothesis to be tested | 5 |
| | Rationale for the selection and coding of data (eg, sound clinical principles or convenience) | NA |
| | Documentation of how data were classified and coded (eg, multiple raters, blinding, and inter-rater reliability) | 5 |
| | Assessment of confounding (e.g. comparability of cases and controls in studies where appropriate) | NA |
| | Assessment of study quality, including blinding of quality assessors; stratification or regression on possible predictors of study results | 5 |
| | Assessment of heterogeneity | NA |
| | Statistical methods (eg, complete description of fixed or random effects models, justification of whether the chosen models account for predictors of study results, dose-response models, or cumulative meta-analysis) in sufficient detail to be replicated | NA |
| *Results* | **Present** | |
| | A graph summarizing individual study estimates and the overall estimate | NA |
| | A table giving descriptive information for each included study | 5 |
| | Results of sensitivity testing (eg, subgroup analysis) | NA |
| | Indication of statistical uncertainty of findings | NA |
| *Discussion* | **Discuss** | |
| | Strengths and weaknesses | 9 |
| | Potential biases in the review process (eg, publication bias) | NA |

| | | |
|---|---|---|
| | Assessment of quality of included studies | 6-7 |
| | Consideration of alternative explanations for observed results | 9 |
| | Generalization of the conclusions (ie, appropriate for the data presented and within the domain of the literature review) | NA |
| | Guidelines for future research | 9 |
| | Disclosure of funding source | 11 |

*Modified from Stroup DF, Berlin JA, Morton SC, Olkin I, Williamson GD, Rennie D, et al. Meta-analysis of observational studies in epidemiology: a proposal for reporting. Meta-analysis Of Observational Studies in Epidemiology (MOOSE) group. JAMA 2000;283:2008–12. Copyrighted © 2000, American Medical Association. All rights reserved.
