## Supplemental Data 2 for "Pattern of Severity of Road Traffic Injuries Among Pedestrians in Low- and Middle-Income Countries: A Systematic Review"

### Additional file 2 – Search Strategy

Initial search covered the keywords:

- ‘Road Traffic Accident/Injury/Crash’
  AND
- ‘Low and Middle-Income Countries’

**PubMed**

*(("road"[All Fields] OR "traffic"[All Fields] OR "transport"[All Fields] OR "transport vesicles"[MeSH Terms] OR "unintentional"[All Fields] OR vehic* OR "motor"[All Fields] OR "motor vehicles"[MeSH Terms]) AND ("accident"[All Fields] OR "accidental injuries"[MeSH Terms] OR "crash"[All Fields] OR "injury"[All Fields] OR "deaths"[All Fields] OR "collision"[All Fields] OR fatal*) AND ("pedestrian"[All Fields] OR "pedestrians"[MeSH Terms] OR "vulnerable road user"[All Fields]) AND (sever* OR ("injury severity score"[MeSH Terms])))*

**Scopus**

*TITLE-ABS-KEY (("road" OR "traffic" OR "transport" OR "transport vesicles" OR "unintentional" OR vehic* OR "motor" OR "motor vehicles") AND ("accident" OR "accidental injuries" OR "crash" OR "injury" OR "deaths" OR "collision" OR fatal*) AND ("pedestrian" OR "pedestrians" OR "vulnerable road user") AND (sever* OR ("injury severity score")))*

**Web of Science**

*ALL = (("road" OR "traffic" OR "transport" OR "transport vesicles" OR "unintentional" OR vehic* OR "motor" OR "motor vehicles") AND ("accident" OR "accidental injuries" OR "crash" OR "injury" OR "deaths" OR "collision" OR fatal*) AND ("pedestrian" OR "pedestrians" OR "vulnerable road user") AND (sever* OR ("injury severity score") ))*

**EMBASE**

*('road'/exp OR 'road' OR 'traffic'/exp OR 'traffic' OR 'transport'/exp OR 'transport' OR 'transport vesicles'/exp OR 'transport vesicles' OR 'unintentional' OR vehic* OR 'motor'/exp OR 'motor' OR 'motor vehicles'/exp OR 'motor vehicles') AND ('accident'/exp OR 'accident' OR 'accidental injuries'/exp OR 'accidental injuries' OR 'crash' OR 'injury'/exp OR 'injury' OR 'deaths' OR 'collision'/exp OR 'collision' OR fatal*) AND ('pedestrian'/exp OR 'pedestrian' OR 'pedestrians'/exp OR 'pedestrians' OR 'vulnerable road user') AND (sever* OR 'injury severity score'/exp OR 'injury severity score')*

*Time filter was applied to restrict our searched articles between 1997 – 2016.
**No restriction was applied for LMICs/ specific country. Authors manually screened for relevant studies of LMICs.
