## Supplemental Data 3 for "Pattern of Severity of Road Traffic Injuries Among Pedestrians in Low- and Middle-Income Countries: A Systematic Review"

**Additional file – 2: World Bank list of low and middle-income countries**

| | | | |
|---|---|---|---|
| Afghanistan | Dominica | Liberia | Sao Tome and Principe |
| Albania | Dominican Republic | Libya | Senegal |
| Algeria | Ecuador | Macedonia, FYR | Serbia |
| American Samoa | Egypt, Arab Rep. | Madagascar | Sierra Leone |
| Angola | El Salvador | Malawi | Solomon Islands |
| Argentina | Equatorial Guinea | Malaysia | Somalia |
| Armenia | Eritrea | Maldives | South Africa |
| Azerbaijan | Ethiopia | Mali | South Sudan |
| Bangladesh | Fiji | Marshall Islands | Sri Lanka |
| Belarus | Gabon | Mauritania | St. Lucia |
| Belize | Gambia, The | Mauritius | St. Vincent and the Grenadines |
| Benin | Georgia | Mexico | Sudan |
| Bhutan | Ghana | Micronesia, Fed. Sts. | Suriname |
| Bolivia | Grenada | Moldova | Swaziland |
| Bosnia and Herzegovina | Guatemala | Mongolia | Syrian Arab Republic |
| Botswana | Guinea | Montenegro | Tajikistan |
| Brazil | Guinea-Bissau | Morocco | Tanzania |
| Bulgaria | Guyana | Mozambique | Thailand |
| Burkina Faso | Haiti | Myanmar | Timor-Leste |
| Burundi | Honduras | Namibia | Togo |
| Cabo Verde | India | Nepal | Tonga |
| Cambodia | Indonesia | Nicaragua | Tunisia |
| Cameroon | Iran, Islamic Rep. | Niger | Turkey |
| Central African Republic | Iraq | Nigeria | Turkmenistan |
| Chad | Jamaica | Pakistan | Tuvalu |
| China | Jordan | Palau | Uganda |
| Colombia | Kazakhstan | Panama | Ukraine |
| Comoros | Kenya | Papua New Guinea | Uzbekistan |
| Congo, Dem. Rep. | Kiribati | Paraguay | Vanuatu |
| Congo, Rep. | Korea, Dem. People’s Rep. | Peru | Venezuela, RB |
| Costa Rica | Kosovo | Philippines | Vietnam |
| Cote d'Ivoire | Kyrgyz Republic | Romania | West Bank and Gaza |
| Cuba | Lao PDR | Russian Federation | Yemen, Rep. |
| Djibouti | Lebanon | Rwanda | Zambia |
| | Lesotho | Samoa | Zimbabwe |

*Accessed in December 2017 from https://data.worldbank.org/income-level/low-and-middle-income
