## Supplemental Data 4 for "Pattern of Severity of Road Traffic Injuries Among Pedestrians in Low- and Middle-Income Countries: A Systematic Review"

**Additional file – 5: List of articles excluded after full-text review**

| Sr No | Author (Year of Publication) | Title | Reason for exclusion |
|---|---|---|---|
| 1 | Joseph et al. (2014) | Epidemiological profile of facial trauma treated at father muller hospital, Thumbay | Data on pedestrian injuries was not available. |
| 2 | Keum (2016) | Analysis of road traffic crashes and injury severity of pedestrian victims in the Gambia | Specific injury severity scoring system was not used. |
| 3 | Yang et al. (2005) | Correlation of different impact conditions to the injury severity of pedestrians in real world accidents | Data on pedestrian injuries was not available. |
| 4 | Porchia et al. (2014) | Effectiveness of two interventions in preventing traffic accidents: a systematic review | Reference list was screened for any relevant and missing articles. No new article was found. |
| 5. | Reith et al. (2015) | Injury pattern, outcome, and characteristics of severely injured pedestrian | Study context was not relevant to LMICs. This information was not available in the title and abstract section. |
| 6. | Lozano et al. (2012) | Global and regional mortality from 235 causes of death for 20 age groups in 1990 and 2010: a systematic analysis for the Global Burden of Disease Study 2010 | Reference list was screened for any relevant and missing articles. |
| 7. | Lateef M (2011) | Spatial patterns monitoring of road traffic injuries in Karachi metropolis | Specific injury severity scoring system was not used. |
| 8. | Olukoga A (2008) | Pattern of road traffic accidents in Durban municipality, South Africa | Specific injury severity scoring system was not used. |
| 9. | Obeng K (2007) | Injury Severity, Vehicle Safety Features, and Intersection Crashes | Study context was not relevant to LMICs. This information was not available in the title and abstract section. |
| 10. | Moe H (2008) | Road traffic injuries among patients who attended the accident and emergency unit of the university of Malaya Medical Centre, Kuala Lumpur | Specific injury severity scoring system was not used. |
| 11. | Adeloye D et al. (2016) | The burden of road traffic crashes, injuries, and deaths in Africa: a systematic review and meta-analysis | Reference list was screened for any relevant and missing articles. |

| | | | |
|---|---|---|---|
| 12. | Mooney S J et al. (2016) | Use of Google Street View to Assess Environmental Contributions to Pedestrian Injury | Specific injury severity scoring system was not used. |
| 13. | Lutge E et al. (2016) | Injury Research: Investigators at University of KwaZulu-Natal Report Findings in Injury Research (A hospital-based surveillance system to assess the burden of trauma in KwaZulu-Natal Province South Africa) | Specific injury severity scoring system was not used. |
| 14. | Zhou M et al. (2016) | Cause-specific mortality for 240 causes in China during 1990-2013: a systematic subnational analysis for the Global Burden of Disease Study 2013 | Reference list was screened for any relevant and missing articles. |
| 15. | Richmond S A et al. (2016) | Trends in unintentional injury mortality in Canadian children 1950-2009 and association with selected population-level interventions | Data on pedestrian injuries was not available. |
| 16. | Hyder A et al. (2016) | The Road Traffic Injuries Research Network: a decade of research capacity strengthening in low- and middle-income countries | Data on pedestrian injuries was not available. |
| 17. | Leidman E et al. (2016) | Road traffic fatalities in selected governorates of Iraq from 2010 to 2013: prospective surveillance | Specific injury severity scoring system was not used. |
| 18. | McClure R et al. (2015) | Contextual Determinants of Childhood Injury: A Systematic Review of Studies with Multilevel Analytic Methods | Reference list was screened for any relevant and missing articles. |
| 19. | Dharmaratne S D et al. (2015) | Road traffic crashes, injury and fatality trends in Sri Lanka: 1938-2013 | Specific injury severity scoring system was not used. |
| 20. | Matzopoulos R et al. (2015) | Injury-related mortality in South Africa: a retrospective descriptive study of post-mortem investigations | Specific injury severity scoring system was not used. |
| 21. | Onywera V O et al. (2013) | Road accidents: a third burden of 'disease' in sub-Saharan Africa | Data on pedestrian injuries was not available. |
| 22. | Verzosa N et al. (2016) | Severity of road crashes involving pedestrians in Metro Manila, Philippines | Specific injury severity scoring system was not used. |

| | | | |
|---|---|---|---|
| 23. | Huang K et al. (2015) | The relationship between injury severity and individual characteristics: a survey in Southern China | Specific injury severity scoring system was not used. |
| 24. | Qiu J et al. (2016) | Analysis of road traffic injuries by road user type in Chongqing, China | Specific injury severity scoring system was not used. |
| 25. | Lutge E et al. (2015) | A hospital-based surveillance system to assess the burden of trauma in KwaZulu-Natal Province South Africa | Specific injury severity scoring system was not used. |
| 26. | McGreevy J et al. (2014) | Road traffic injuries in Yaounde, Cameroon: A hospital-based pilot surveillance study | Specific injury severity scoring system was not used. |
| 27. | Bumbasirevic M et al. (2014) | Severe road traffic injuries and youth: a 4-year analysis for the city of Belgrade | Data on pedestrian injuries was not available. |
| 28. | Kamruzzaman M et al. (2014) | Analysis of Traffic Injury Severity in Dhaka, Bangladesh | Data on pedestrian injuries was not available. |
| 29. | Parkinson F et al. (2013) | Road traffic crashes in South Africa: The burden of injury to a regional trauma centre | Specific injury severity scoring system was not used. |
| 30. | Parkinson F et al. (2013) | Patterns of injury seen in road crash victims in a South African trauma centre | Specific injury severity scoring system was not used. |
| 31. | Minca D G et al. (2013) | Profile of persons involved in traffic accidents in Romania | Data on pedestrian injuries was not available. |
| 32. | Zhao H et al. (2013) | An Investigation on the Head Injuries of Adult Pedestrians by Passenger Cars in China | Specific injury severity scoring system was not used. |
| 33. | Velloso et al. (2012) | On-the-spot study of pedestrian crashes on Brazilian Federal District rural highways crossing urban areas | Specific injury severity scoring system was not used. |
| 34. | Pernica J F et al. (2011) | Risk factors predisposing to pedestrian road traffic injury in children living in Lima, Peru: a case-control study | Specific injury severity scoring system was not used. |
| 35. | Zimmerman K et al. (2011) | Road traffic injury incidence and crash characteristics in Dar es Salaam: A population-based study | Data on pedestrian injuries was not available. |
| 36. | Guerrero A et al. (2010) | Paediatric road traffic injuries in urban Ghana: a population-based study | Specific injury severity scoring system was not used. |
| 37. | Malta D C et al. (2011) | Analysis of the occurrence of traffic injuries and related factors according to the | Specific injury severity scoring system was not used. |

| | | | |
|---|---|---|---|
| | | National Household Sample Survey (PNAD) - Brazil, 2008 | |
| 38. | Dandona R et al. (2011) | Road use pattern and risk factors for non-fatal road traffic injuries among children in urban India | Specific injury severity scoring system was not used. |
| 39. | Li F et al. (2010) | A study of head brain injuries in car-to-pedestrian crashes with reconstructions using in-depth accident data in China | Specific injury severity scoring system was not used. |
| 40. | Zhao H et al. (2010) | Investigation of 184 passenger car-pedestrian accidents | Specific injury severity scoring system was not used. |
| 41. | Bhalla K et al. (2009) | Building national estimates of the burden of road traffic injuries in developing countries from all available data sources: Iran | Data on pedestrian injuries was not available. |
| 42. | Dandona R et al (2008) | Incidence and burden of road traffic injuries in urban India | Specific injury severity scoring system was not used. |
| 43. | Adesunkanmi A R et al. (2000) | Road traffic accidents to African children: assessment of severity using the Injury Severity Score (ISS) | Data on pedestrian injuries was not available. |
| 44. | Al-Omari B H et al. (2013) | Analysis of Pedestrian Accidents in Irbid City, Jordan | Specific injury severity scoring system was not used. |
| 45. | Nie J et al. (2010) | A Study on Pedestrian Injuries based on Minivan and Sedan Real-World Accidents | Specific injury severity scoring system was not used. |
| 46. | Siram S M et al. (2010) | Does the Pattern of Injury in Elderly Pedestrian Trauma Mirror That of The Younger Pedestrian? | Specific injury severity scoring system was not used. |
| 47. | Derry J D et al. (2010) | Pedestrians Injury Patterns in Ghana | Specific injury severity scoring system was not used. |
