## Supplemental Data 5 for "Pattern of Severity of Road Traffic Injuries Among Pedestrians in Low- and Middle-Income Countries: A Systematic Review"

**Additional file – 5: Modified Newcastle-Ottawa Scale adapted for cross-sectional studies**

**Selection: (Maximum 5 stars)**

**1) Representativeness of the sample: (Maximum 1 star)**

a) Truly representative of the average in the target population. * (all subjects or random sampling)
b) Somewhat representative of the average in the target population. * (non-random sampling)
c) Selected group of users.
d) No description of the sampling strategy.

**2) Sample size: (Maximum 1 star)**

a) Justified and satisfactory. * [Sample size calculated, and it was adequate according to reviewer]
b) Not justified. [Rest all]
c) No description of the sample size calculation. [in cases of large multi-centric studies, it may be classified as (a)]

**3) Non-respondents: (Maximum 1 star)**

a) Comparability between respondents and non-respondent's characteristics is established, and the response rate is satisfactory. *
b) The response rate is unsatisfactory, or the comparability between respondents and non-respondents is unsatisfactory.
c) No description of the response rate or the characteristics of the responders and the non-responders.

**4) Ascertainment of exposure: (Maximum 2 star)**

a) Secured record (e.g. Hospital/Registry) **
b) Structured interview or questionnaire *
c) Written self-report
d) No description

**Comparability: (Maximum 2 stars)**

**1) The subjects in different outcome groups are comparable, based on the study design or analysis. Confounding factors are controlled.**

a) The study controls for the most important factor (Age Group). *
b) The study control for any additional factor. * (Gender, Residence-Urban/Rural)

**Outcome: (Maximum 3 stars)**

**1) Assessment of the outcome: (Maximum 2 stars)**

a) Validated measurement tool. **
b) Non-validated measurement tool, but the tool is available or described. *
c) No description of the measurement tool.

**2) Statistical test: (Maximum 1 stars)**

a) The statistical test used to analyze the data is clearly described and appropriate, and the measurement of the association is presented, including confidence intervals and the probability level (p value). *

b) The statistical test is not appropriate, not described, or incomplete.

Most important factor for comparability is the age group which is an important increasing risk factor of RTI and mortality in young age groups. Thus, the principal factors were identified for each study. Few commonly selected factors were sex, residence – urban/rural.
