## Supplemental Data 6 for "Pattern of Severity of Road Traffic Injuries Among Pedestrians in Low- and Middle-Income Countries: A Systematic Review"

### Additional File 6: Summary of studies reported severity pattern of road traffic injuries among pedestrians in LMICs

| **Authors-Year of Publication** | **Study location, study setting and study design, sampling method** | **Severity scale** | **Participants characteristics** (type of RTIs; sample size; sex and distribution) | **Salient features** |
|---|---|---|---|---|
| **NieJin et al-2010** | China; population-based cross-sectional study | AIS | Crashes with car and mini vans (79 sedans; 31 minivans)<br>Sample size: 110<br>All age groups | 1. Severe injuries (AIS 3+) due to crash with sedans were 35% and due to minivans were 50.5%.<br>2. Minivan poses a greater risk to the pedestrian than sedan at the same impact speed.<br>3. A significant correlation between the impact speed and severity of pedestrian injuries was documented. There is a significant relationship between pedestrian head injuries (AIS 3+) and injury parameters (head injury cases, head linear acceleration, head angle acceleration, angle velocity). |
| **Zhao et al-2013** | Beijing, Shanxi Province, and Chongqing, China: population-based cross-sectional study | AIS | Crashes with car<br>Sample size: 285<br>Aged 16 years and above | 1. 10.9% (31) were aged 16-25 years, 31.6% (90) were aged 26-45 years, 31.2% (89) aged 46-60 years, and 26.3% (75) were aged 60 years and above.<br>2. Based on AIS for head injuries, 38% had AIS 3-4, 21% had AIS 1-2, and 19% had AIS 5-6.<br>3. Crossing the road was a major cause responsible for 78% (222) injuries, followed by walking along the road 17% (48), and standing still 5% (15).<br>4. A higher pedestrian fatality risk was associated with age over 46 years, impact speed over 40 km/h, and a higher likelihood of the victim’s head striking the windscreen frame/A-pillar and of the victim sustaining a head injury. |

| **Authors-Year of Publication** | **Study location, study setting and study design, sampling method** | **Severity scale** | **Participants characteristics** (type of RTIs; sample size; sex and distribution) | **Salient features** |
|---|---|---|---|---|
| **NieJin et al-2010** | China; population-based cross-sectional study | AIS | Crashes with car and mini vans (79 sedans; 31 minivans)<br><br>Sample size: 110<br><br>All age groups | 1. Severe injuries (AIS 3+) due to crash with sedans were 35% and due to minivans were 50.5%.<br>2. Minivan poses a greater risk to the pedestrian than sedan at the same impact speed.<br>3. A significant correlation between the impact speed and severity of pedestrian injuries was documented. There is a significant relationship between pedestrian head injuries (AIS 3+) and injury parameters (head injury cases, head linear acceleration, head angle acceleration, angle velocity). |
| **Aladelusi et al - 2014** | Ibadan, Nigeria: hospital-based cross-sectional study, prospective recruitment of all eligible participants | MFISS | RTIs involving maxillofacial injuries<br><br>Sample size: 46 (males 52% and females 48%)<br><br>All age groups | 1. 22.9% participants were pedestrians which are predominantly in the age group 11-20 years (21%), followed by 0-10 years and 21-30 years and 31-40 years (17%, each). After that, it has decreased significantly.<br>2. Majority of RTI victims (65.2%) were hit by a car or minibus, followed by motorcycles (30.4%), and trucks (4.4%).<br>3. The most severe MFISS was observed in the age group 21-30 years, while the least severe injury observed was in the 71 – 80 years age group.<br>4. The most severe maxillofacial injury was seen in the victim of car/minibus pedestrian crashes 46% (21); 17 per cent (8) had a fatal outcome.<br>5. The median MFISS score of victims of car/minibus was 9.0, and motorcycle collision was 4.0.<br>6. Soft tissue injury was the most common maxillofacial injury 50% (23) and head injury was the commonest concomitant injury 66.7% (12) observed. |

| Authors-Year of Publication | Study location, study setting and study design, sampling method | Severity scale | Participants characteristics (type of RTIs; sample size; sex and distribution) | Salient features |
|---|---|---|---|---|
| **NieJin et al-2010** | China; population-based cross-sectional study | AIS | Crashes with car and mini vans (79 sedans; 31 minivans)<br>Sample size: 110<br>All age groups | 1. Severe injuries (AIS 3+) due to crash with sedans were 35% and due to minivans were 50.5%.<br>2. Minivan poses a greater risk to the pedestrian than sedan at the same impact speed.<br>3. A significant correlation between the impact speed and severity of pedestrian injuries was documented. There is a significant relationship between pedestrian head injuries (AIS 3+) and injury parameters (head injury cases, head linear acceleration, head angle acceleration, angle velocity). |
| **Solagberu et al-2014** | Ikeja, Nigeria: population-based cross-sectional study, prospective recruitment of all eligible participants | GCS | All RTIs<br>Sample size: 226 (males - 114 and females - 112)<br>Children aged up-to 15 years | 1. 18.5% (42) were aged 0-4 years, 40.3% (91) aged 5–9 years, and 41.1% (93) aged 10–15 years.<br>2. They had collision with car (36.7%, 83), motorcycles (33.6%, 76), buses (18.1%, 41), others (6.6%, 15), and 11 undetermined vehicles.<br>3. Crossing the road was a major cause responsible for 74.3% (168) injuries, followed by hit at the bus stop 8.4% (19), walking along with the pavement 9.7% (22), and standing in front of house/shop 7.5% (17).<br>4. Anatomical regions injured were head (42.9%), lower limbs (35.4%) and upper limb injury 9.7% (22).<br>5. Fatality risks were truck collisions (OR 5.97, 95% CI 1.01-34.98), female child (OR 4.25, 95% CI 1.36-13.24), head injury (OR 4.18, 95% CI 1.45-12.05) and age up to 4 years (OR 3.7, 95% CI 1.38-9.88).<br>6. According to GCS, 37% (40) subjects were mild, 30.5% (33) moderate, and 32.4% (35) were severe head injuries (among total 108 head injuries). Among which crossing the road produced 75%, 81%, and 85% mild, moderate, and severe head injuries, respectively. |

| **Authors-Year of Publication** | **Study location, study setting and study design, sampling method** | **Severity scale** | **Participants characteristics** (type of RTIs; sample size; sex and distribution) | **Salient features** |
|---|---|---|---|---|
| **NieJin et al-2010** | China; population-based cross-sectional study | AIS | Crashes with car and mini vans (79 sedans; 31 minivans)<br>Sample size: 110<br>All age groups | 1. Severe injuries (AIS 3+) due to crash with sedans were 35% and due to minivans were 50.5%.<br>2. Minivan poses a greater risk to the pedestrian than sedan at the same impact speed.<br>3. A significant correlation between the impact speed and severity of pedestrian injuries was documented. There is a significant relationship between pedestrian head injuries (AIS 3+) and injury parameters (head injury cases, head linear acceleration, head angle acceleration, angle velocity). |
| **Tokdemir et al-2009** | Firat, Turkey: hospital-based, secondary data analysis of hospital records | GCS | RTIs involving head injuries<br>Sample size: 233 (males - 163, females - 70)<br>All age groups | 1. The pediatric (≤16 years: 65.4%) and elderly (≥65 years: 64.7%) groups were frequently involved as pedestrians in motor vehicle accidents; adults 17–64 years of age were involved as pedestrians at a lower rate (25.4%, p<0.001).<br>2. According to GCS, 59.7 % (139) subjects were mild, 15.9 % (37) moderate, and 24.5 % (57) were severe head injuries.<br>3. Mortality rate among them was 18.3% (43). |

[#] If age not given, we considered all age groups.
