## Supplemental Data 7 for "Pattern of Severity of Road Traffic Injuries Among Pedestrians in Low- and Middle-Income Countries: A Systematic Review"

**Additional file - 7: Quality Appraisal of shortlisted studies with Newcastle-Ottawa Scale**

| Authors-Year of Publication | Representativeness of the sample | Sample size | Non-respondents | Ascertainment of the exposure | Comparability | Assessment of the outcome | Statistical test | Total |
|---|---|---|---|---|---|---|---|---|
| **Aladelusi et al -2014** | 0 | 0 | 0 | 0 | 2 | 2 | 0 | 4 |
| **NieJin et al-2010** | 0 | 0 | 0 | 2 | 1 | 2 | 1 | 6 |
| **Solagberu et al-2014** | 0 | 0 | 0 | 0 | 1# | 2 | 1 | 4 |
| **Tokdemir et al-2009** | 0 | 0 | NA* | 2 | 1 + | 2 | 1 | 6 |
| **Zhao et al-2013** | 0 | 0 | 0 | 1 | 2 | 2 | 1 | 6 |

**NA- Not applicable due to limitation (secondary data), #Children up to 15 years were included, +Adolescence above 16 years were selected.*
